# Epidemiological and Clinical Features of Mpox during the Clade Ib Outbreak in South-Kivu, Democratic Republic of the Congo: a Prospective Cohort Study

**DOI:** 10.1101/2024.11.18.24316975

**Authors:** Isabel Brosius, Emmanuel Hasivirwe Vakaniaki, Guy Mukari, Papy Munganga, Jean Claude Tshomba, Elise De Vos, Eugene Bangwen, Yves Mujula, Achilleas Tsoumanis, Christophe Van Dijck, Aimé Alengo Odud, Léandre Mutimbwa-Mambo, Franklin Mweshi Kumbana, Jenestin Babingwa Muunga, Divin Mazambi Mambo, Jems Wakilongo Zangilwa, Steeven Bilembo Kitwanda, Sarah Houben, Nicole A. Hoff, Jean-Claude Makangara-Cigolo, Eddy Kinganda-Lusamaki, Martine Peeters, Anne W. Rimoin, Jason Kindrachuk, Nicola Low, Patrick DMC Katoto, Espoir Bwenge Malembaka, John H. Amuasi, Olivier Tshiani-Mbaya, Dally Muamba Kambaji, Richard Kojan, Cris Kacita, Daniel Mukadi-Bamuleka, Steve Ahuka-Mundeke, Koen Vercauteren, Tony Wawina-Bokalanga, Jean-Jacques Muyembe-Tamfum, Sabin Sabiti Nundu, Laurens Liesenborghs, Placide Mbala-Kingebeni

## Abstract

**Background:** Clade Ib, a new strain of the Clade I monkeypox virus, emerged in Eastern Democratic Republic of the Congo, sparking an international outbreak. Comprehensive studies are needed to assess its transmission dynamics and clinical presentation.

**Methods:** We conducted a prospective observational cohort study at Kamituga General Hospital in South Kivu, DRC, between May 2 and October 9, 2024. Sociodemographic, exposure and clinical data were collected from mpox suspected cases. Cases confirmed by Xpert® Mpox PCR were presumed Clade Ib infections (awaiting Clade confirmation) and followed through hospitalization and on days 29 and 59 post-diagnosis to assess clinical progression and outcomes.

**Findings:** Of 511 included suspected cases, 431 (84%) tested PCR positive; with 205 being women (47%). Age distribution was bimodal, with 279 (65%) individuals aged 15-34 years, and 63 (15%) children under five. Most cases (59%) reported contact with a suspected or confirmed mpox case; among adults, this was primarily a spouse, colleague or sexual partner, while for children, the primary contacts were parents or siblings. Comorbidities were rare (4%), including six (1%) HIV infections. Prodromal symptoms were present in 346 (88%) patients, active skin lesions in 414 (96%), mucosal lesions in 338 (82%), and lymphadenopathy in 295 (71%). In adults, lesions were more concentrated in the genital area, with 90% of adults presenting lesions in this region. In contrast, only 39% of children had genital lesions, with lesions more frequently found elsewhere on the body. Among 427 hospitalized patients, two deaths (0·5%) occurred. Among 315 patients with detailed hospital follow-up, complications were primarily genito-urinary (55%) or cutaneous (40%). Four of six pregnant women with recorded outcome (67%) had adverse pregnancy outcomes. Significant sequelae at days 29 and 59 were rare.

**Interpretation:** Clade Ib MPXV infections presented differently from previously reported Clade Ia and Clade IIb infections. In adults, the disease primarily affected the genito-urinary system, compatible with sexual transmission, while children mostly manifested extragenital lesions.

**Funding:** European & Developing Countries Clinical Trials Partnership (EDCTP2 and EDCTP3); Belgian Directorate-General Development Cooperation and Humanitarian Aid; Research Foundation – Flanders

## Introduction

Mpox is a disease caused by monkeypox virus (MPXV). Until recently, human mpox was considered an endemic zoonosis, linked to contact with wildlife infected with MPXV Clade I in Central Africa or MPXV Clade II in West Africa.^1^ However, in recent years, two distinct lineages of MPXV capable of sustained human-to-human transmission have emerged, leading to major international outbreaks.^2^

In 2017, a subclade of Clade II, named Clade IIb began to circulate in Nigeria through human-to-human transmission, including through sexual contact.^3^ The subsequent international spread of Clade IIb culminated in the 2022 global outbreak, with over 100,000 documented mpox cases to date. The outbreak overwhelmingly affected adult men – often identifying as men-who-have-sex-with-men (MSM) –, who predominantly presented with relatively mild disease and predominantly anogenital or oral lesions.^1,4^ The overall case fatality rate (CFR) was below 0·1%.^2^

A similar scenario is currently unfolding with Clade I in the Democratic Republic of the Congo (DRC). The country notified a substantial increase in Clade I MPXV infections over multiple decades, primarily in remote rural areas, where outbreaks are largely due to zoonotic spillover and predominantly impact children.^5,6^ Historically, Clade I infections have been associated with a generalized papulopustular rash, with potentially severe outcomes and a reported CFR of 2·9% among suspect cases.^5,7,8^

In September 2023, however, a new lineage of Clade I MPXV emerged in a densely populated urban and previously unaffected region in South Kivu, DRC. ^9–11^ Phylogenomically, the lineage constitutes a distinct cluster, named Clade Ib, distinct from other Clade I lineages that are now denoted Clade Ia.^10^ The subsequent rapid human-to-human spread of Clade Ib in South Kivu resulted in the notification of 2,672 confirmed mpox cases between January 1 and October 20, 2024. Of these, 157 samples were sequenced, with 156 showing Clade Ib.^12,13^ From South Kivu, Clade Ib spread to other provinces within the country and cross-border to neighboring countries. Consequently, in August 2024, both the Africa Center for Disease Control (CDC) and the World Health Organization (WHO) issued emergency declarations.^14^

Unlike Clade Ia, early reports about Clade Ib mpox suggested that mainly adolescents and young adults were affected, with lower disease severity and predominantly genital lesions.^10,11,13^ Clade Ib, therefore, exhibits notable similarities with Clade IIb mpox. However, robust clinical data are lacking.

Large prospective studies are needed to characterize the ongoing Clade Ib mpox outbreak, given apparent changes in affected populations, clinical presentation, disease outcome, and transmission dynamics. This manuscript describes the epidemiological and clinical characteristics of the Clade Ib mpox outbreak in South Kivu, DRC.

## Material and Methods

### Study design, Site and Participants

The Mpox Biology, Outcome, Transmission, and Epidemiology - Kamituga (MBOTE - Kamituga) study is a prospective observational cohort study of Clade Ib MPXV-infected individuals (clinicaltrials.gov: NCT06652646). Participants were enrolled at Kamituga General Hospital (KGH), which hosts a large mpox treatment center for Kamituga and surrounding health zones. Kamituga Health Zone, the initial epicenter of the Clade Ib mpox outbreak, is an urbanized region in South-Kivu province, eastern DRC, with a population estimated at 242,000 people.^15^ On-site diagnostic testing capacity for mpox was installed, and a new isolation unit was set up with an admission capacity of 37 beds.

All individuals suspected of having mpox who presented to the hospital for diagnostic testing and care, and who provided written informed consent were included in the study. Participants were enrolled prior to diagnostic testing and underwent a baseline investigation. Only individuals who tested positive via Mpox Xpert® PCR were subsequently followed up longitudinally.

### Procedures and Data collection

A baseline standardized case report form (CRF), programmed into a REDCap™ database (Vanderbilt University, Nashville, TN), was completed by a study physician. The CRF captured data on exposure, behaviors, socio-demographic profile, medical history including HIV and smallpox vaccination status, as well as presence and onset date of symptoms. Questions about sexual behaviors were restricted to people over 12 years old. Physical examination findings at enrolment included lesion count, lymphadenopathy, and presence or absence of complications.

Skin lesions and oropharyngeal swabs were taken for on-site diagnostic testing with Xpert® Mpox cartridges on the 4-module GeneXpert system (Cepheid, Sunnyvale, California CA, USA). This automated real-time PCR detects non-variola Orthopoxvirus (OPXV) and MPXV Clade II DNA .^16^ A sample was considered positive for MPXV Clade I, if non-variola OPXV DNA was detected with a cycle threshold value below 39 and MPXV Clade II PCR was negative, following guidelines from the DRC laboratory working group led by WHO. At the time of preprint publication, Clade Ib confirmation is ongoing.

Individuals with a positive lesion or oropharyngeal sample were admitted to KGH for isolation and medical care and followed-up by the study team. The evolution of symptoms, signs, and complications was documented in a CRF. Patients received supportive care, consisting of symptomatic therapy (analgesia, antipyretics), nutritional support, skin care, correction of hypovolemia, antibacterial and antimalarial treatment where indicated. No MPXV-specific antivirals were available. Malaria and HIV rapid diagnostic tests, hemoglobin assays, point-of-care glucometry, and HCG dipstick tests were performed at the discretion of the treating physician. Testing for other sexually transmitted infections, Varicella, bacterial infections, and biochemical analyses were not available. Patients were discharged at the discretion of the treating physician team.

After discharge, participants were invited for follow-up visits on days 29 and 59 post-mpox diagnosis to record data on residual or new signs and symptoms. Pregnant women were advised to return to KGH for any pregnancy-related complications, or at the start of labor. Pregnancy related outcomes were captured in a dedicated CRF.

### Statistical analysis

Demographic, clinical and behavioral characteristics were described using medians and interquartile ranges (IQR) for continuous variables and counts and percentages for categorical variables. For the representation of the clinical data, participants were categorized into three age groups based on expected physiological and exposure differences: children under five years, children aged five to 14 years, and individuals aged 15 years and older. The latter group will be referred to as adults throughout the manuscript. The association between sociodemographic and exposure characteristics and MPXV-PCR test results among mpox suspected cases was tested using Fisher exact test for categorical characteristics and Mann-Whitney non-parametric test for continuous variables. The resulting p-values were corrected for multiple testing using the Benjamini-Hochberg method. The time from symptom and lesion onset to resolution were calculated and the results were presented using survival Kaplan-Meier curves. Data were analyzed using R statistical software, version 4.3.1.

### Ethical considerations

The study was approved by the Ethics Committee of the University of Kinshasa (approval ID ESP/CE/78/2024) and the Ethics Committee of the University Hospital Antwerp (ID 6383). Written consent was obtained prior to all study procedures from participants aged 18 years and older, and from parents or guardians for minors. Children from 12 to 17 years old received age-appropriate information and signed an assent form. Pictures for publication were taken with additional written consent. All participants received reimbursement of transportation costs for visits and free care and nutrition.

## Results

Between May 2 and October 9, 2024, 511 people with suspected mpox were enrolled in the study (Supplementary figure 1) of which 431 (84%) tested positive on Xpert® PCR.

### Demographics, Exposure and Risk Factors

Of all 431 confirmed cases, 205 (47·6%) were women (Table 1). The median age was 22 (IQR 17-29, range 1 week to 79 years). Individuals aged 15-34 accounted for 65% of all infections, while children aged 10-14, 5-9, and under five years represented 3%, 4%, and 15%, respectively. (Figure 1A) The most reported occupations were mine worker (100, 24%) and sex worker (53, 13%).

**Table 1:** Demographic profile and exposure risk of suspect mpox cases in Kamituga, DRC, by infection status

|  | All suspected cases<br>(n = 511) | MPXV positive<br>(n = 431) | MPXV negative<br>(n = 80) | Adjusted<br>p value |
| --- | --- | --- | --- | --- |
| **Demographics** | .. | .. | .. |  |
| Age, years - median (IQR) | 22 (14-29) | 22 (17-29) | 17 (4-31) | 0.034 |
| Age group – n (%) | .. | .. | .. | 0.005 |
| 0-4 years | 84/511 (16%) | 63/431 (15%) | 21/80 (26%) | .. |
| 5-9 years | 24/511 (5%) | 18/431 (4%) | 6/80 (8%) | .. |
| 10-14 years | 22/511 (4%) | 14/431 (3%) | 8/80 (10%) | .. |
| 15-34 years | 311/511 (61%) | 279/431 (65%) | 32/80 (40%) | .. |
| 35-49 years | 58/511 (11%) | 52/431 (12%) | 6/80 (8%) | .. |
| ≥ 50 years | 12/511 (2%) | 5/431 (1%) | 7/80 (9%) | .. |
| Female – n (%) | 241/511 (47%) | 205/431 (47%) | 36/80 (45%) | 0.871 |
| Occupation – n (%) | .. | .. | .. | 0.005 |
| Sex worker | 57/498 (11%) | 53/419 (13%) | 4/79 (5%) | .. |
| Mine worker | 106/498 (21%) | 100/419 (24%) | 6/79 (8%) | .. |
| Healthcare worker | 4/498 (1%) | 4/419 (1%) | 0/79 (0%) | .. |
| Barman | 6/498 (1%) | 6/419 (1%) | 0/79 (0%) | .. |
| Teacher | 3/498 (1%) | 2/419 (1%) | 1/79 (1%) | .. |
| Student | 53/498 (11%) | 40/419 (2%) | 13/79 (17%) | .. |
| Services | 87/498 (18%) | 70/419 (17%) | 8/79 (10%) | .. |
| Manual worker | 42/498 (8%) | 35/419 (8%) | 7/79 (9%) | .. |
| Unemployed | 6/498 (1%) | 2/419 (1%) | 4/79 (5%) | .. |
| **Exposure** | .. | .. | .. | .. |
| Exposure to animal reservoir<sup>£</sup> – n (%) | 63/441 (14%) | 58/366 (16%) | 5/75 (7%) | 0.105 |
| Sex in the previous 3 weeks – n (%) | 324/402 (81%) | 293/348 (84%) | 31/54 (57%) | 0.001 |
| # of partners – median (IQR) | 1 (1-3) | 1 (1-3) | 1 (1-1) | 0.055 |
| Engaged in transactional sex | 143/369 (39%) | 135/318 (42%) | 8/51 (16%) | 0.003 |
| Consistent use of condoms<sup>\$</sup> | 13/311 (4%) | 12/280 (4%) | 1/31 (3%) | 1.000 |
| Contact with suspected or confirmed case – n (%) | 286/510 (56%) | 253/430 (59%) | 33/80 (41%) | 0.015 |
| contacts per participant, n – median (IQR) | 1 (1-2) | 1 (1-2) | 1 (1-2) | 1.000 |
| Multiple or continuous exposure | 255/270 (94%) | 225/239 (94%) | 30/31 (97%) | 1.000 |
| Time since last exposure, days – median (IQR) | 4 (1-10) | 4 (1-10) | 1 (1-5) | 0.006 |
| Relation with the contact<sup>#</sup> – n (%) | .. | .. | .. | 0.020 |
| Family (same household) | 144/279 (52%) | 121/247 (49%) | 23/32 (72%) | .. |
| Family (different household) | 12/279 (4%) | 7/247 (3%) | 5/32 (16%) | .. |
| Neighbor | 19/279 (7%) | 17/247 (7%) | 2/32 (6%) | .. |
| Colleague | 88/279 (32%) | 81/247 (33%) | 7/32 (22%) | .. |
| Sexual | 50/279 (18%) | 50/247 (20%) | 0/32 (0%) | .. |
| Sexual partner | 11/279 (4%) | 11/247 (5%) | 0/32 (0%) | .. |
| Client of sex worker | 17/279 (6%) | 17/247 (7%) | 0/32 (0%) | .. |
| Sex worker | 21/279 (8%) | 21/247 (9%) | 0/32 (0%) | .. |
| Patient of the participant | 2/279 (1%) | 2/247 (1%) | 0/32 (0%) | .. |
| Other | 5/279 (2%) | 5/247 (2%) | 0/32 (0%) | .. |
| Type of exposure§ – n (%) | .. | .. | .. | .. |
| Prolonged (> 15 min) discussion | 274/286 (96%) | 243/253 (96%) | 31/33 (94%) | 0.809 |
| Direct skin to skin contact | 279/286 (98%) | 246/253 (97%) | 33/33 (100%) | 1.000 |
| Sexual contact | 114/286 (40%) | 107/253 (42%) | 7/33 (21%) | 0.058 |
| Care giving | 67/286 (23%) | 57/253 (23%) | 10/33 (30%) | 0.562 |
| Sharing meals and/or eating utensils | 254/286 (89%) | 227/253 (90%) | 27/33 (82%) | 0.363 |
| Sharing bed and/or bedlinen | 197/286 (69%) | 176/253 (70%) | 21/33 (64%) | 0.733 |
| Sharing clothes | 36/286 (13%) | 29/253 (12%) | 7/33 (21%) | 0.258 |

Data are median and IQR for numeric variables and n/N (%) for categorical variables. N signifies the number of included individuals with available data. <sup>‡</sup>Exposure to animal reservoir is defined as involvement in activities such as hunting, capturing, or consuming rodents, eating bushmeat, or handling or consuming animals found dead.
<sup>§</sup>Consistent use of condoms means the use of condoms for (nearly) all sexual contact. <sup>#</sup>Spouses were reported as family members. The sum exceeds 100% because the relationship is defined per index case, 1 participant can have > 1 index case. <sup>§</sup>Multiple answers per participant are possible, the numerator indicates the number of participants that had the specific exposure type with at least one index case.
Abbreviations: IQR = interquartile range, min = minutes, MPXV = monkeypox virus

**Figure 1.**
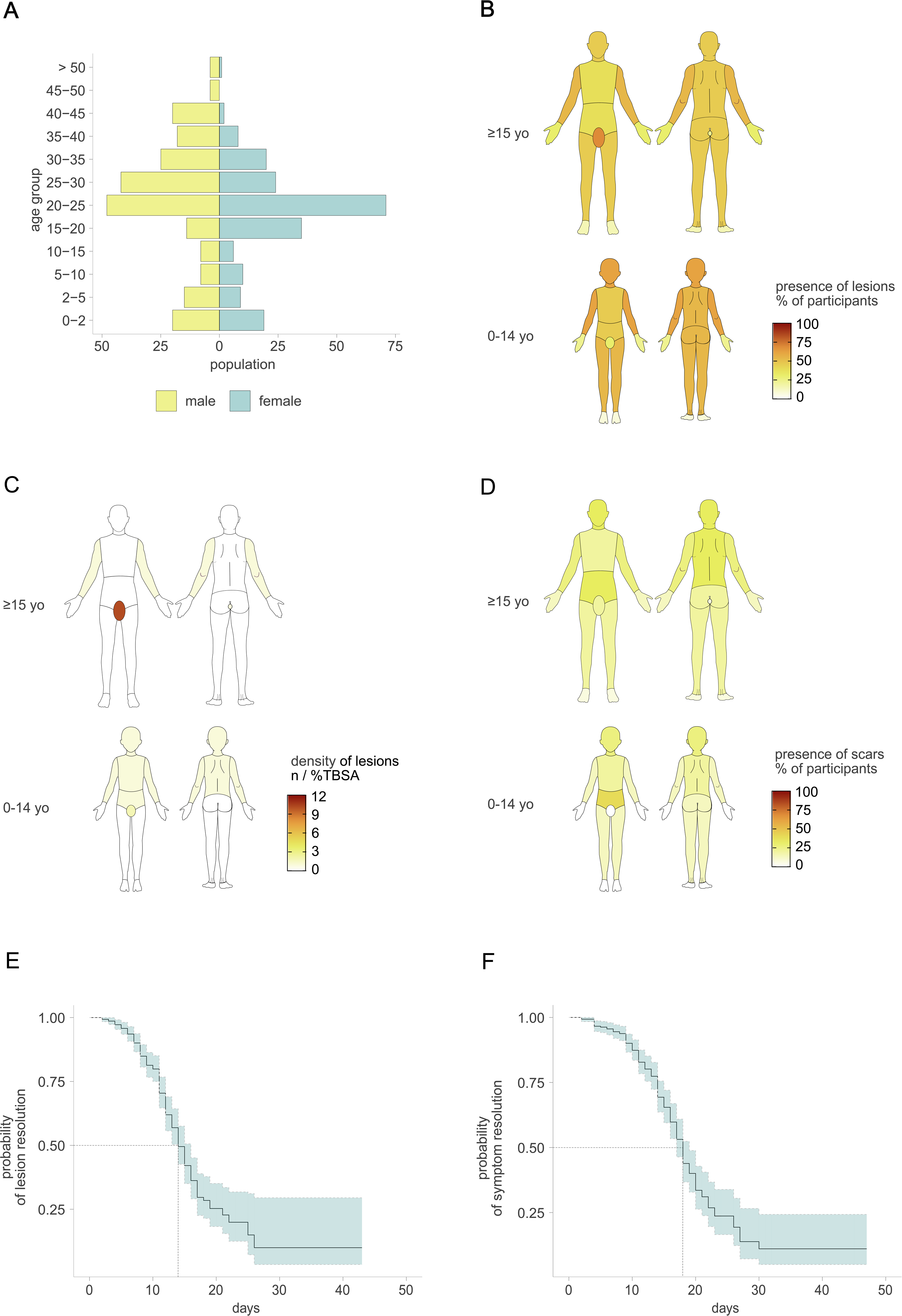
Demographics, Clinical Presentation and Evolution of Clade Ib mpox, South-Kivu, DRC. (A) Age and sex distribution of confirmed mpox participants, (B) Body map indicating the proportion of participants with lesions in a given body region at inclusion for adults (>= 15 years) and children 0-14 years. (C) Body map indicating density of skin lesions per body region for individuals of 15 years old and above, and children aged 0 to 14 years old, expressed as mean number of lesions (n) per percentage of total body surface area (TBSA) according to a modified Lund-Browder chart for estimated TBSA. (D) Body map indicating the proportion of participants with scars in a given body region on day 59 after diagnosis for adults (>= 15 years) and children 0-14 years. (E) Kaplan-Meier estimates for time from onset of active lesions to resolution, with 95% confidence interval. (F) Kaplan-Meier estimates for time from onset of symptoms to resolution of all symptoms, with 95% confidence Interval.

**Figure 2.**
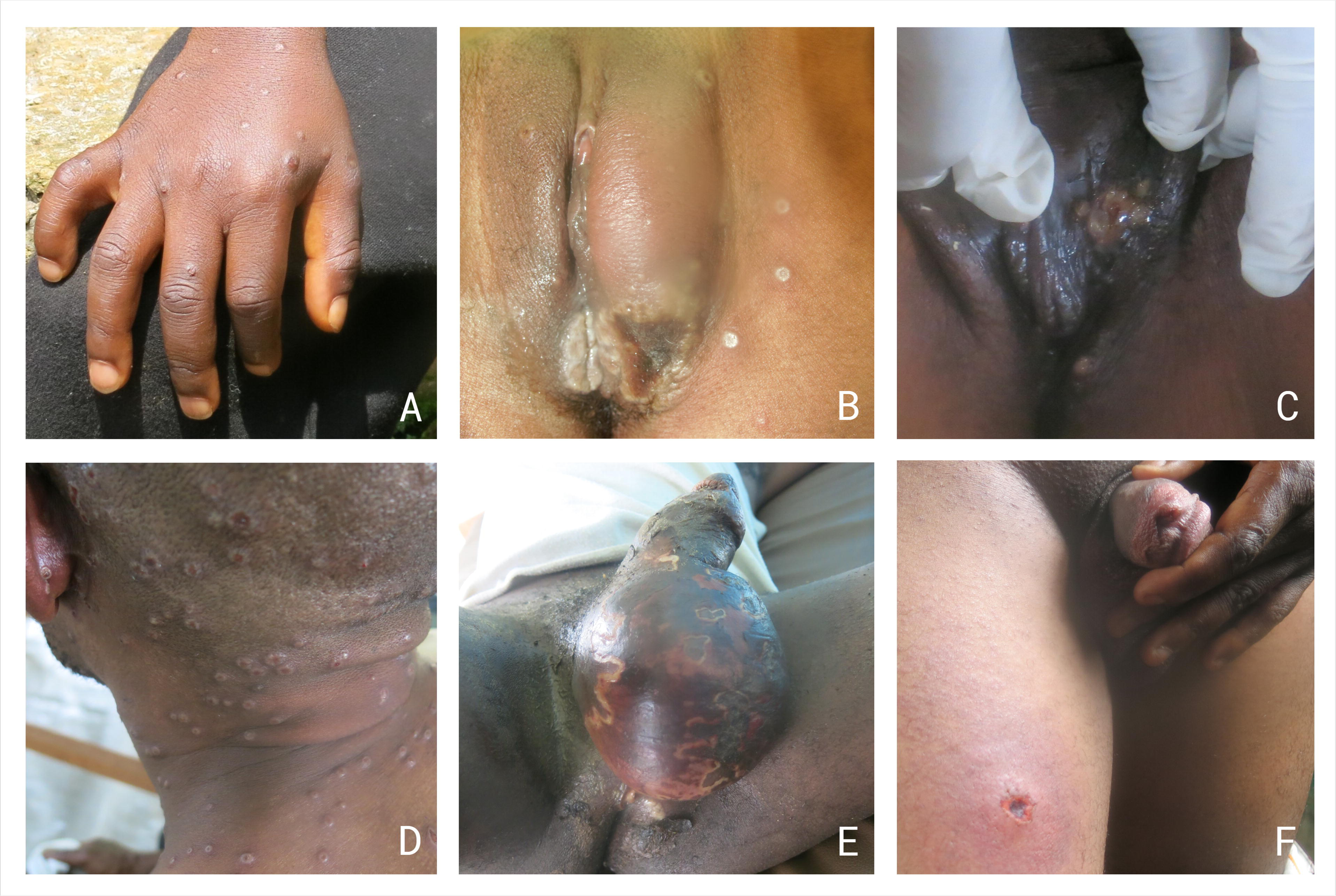
Clinical images of skin and genital mpox lesions. (A) Papular, umbilicated and ulcerated lesions on the hand. (B)Complex skin and genital mucosal lesion with labial edema, surrounded by papules and umbilicated genital skin lesions in adult female. (C) Complex genital skin lesion in adult female. (D) Umbilicated lesions with high lesion density on the head and neck. (E) Genital mpox complicated with Fourniers gangrene in an adult male. (F) Ulcerated and necrotic lesions on the penis and upper thigh in adult male.

Among confirmed cases, 59% (n=253) reported contact with one or more suspected or confirmed mpox cases in the three weeks before diagnosis. Among the 57 children with this exposure, potential infectors were primarily household members (n=75, 83%). These household contacts were predominantly parents (47%) or siblings (53%). Similarly, the 197 adults with known contact, most frequently reported household members (n=91, 37%) as potential infectors, with spouses being the most common household contact (56/91, 62%). Other frequently mentioned potential infectors among adults were colleagues (n=81, 33%) and sexual partners outside the household (n=50, 21%). The most common exposure types were skin-to-skin contact (n=246, 97%), prolonged conversation (>15 min) (n=243, 96%), shared meals or utensils (n=227, 90%), and shared bedding (n=176, 70%), with 42% (n=107) reporting sexual exposure.

Of the 348 confirmed mpox patients aged 12 years or older questioned on sexual behavior, 293 (84%) reported having sex in the previous three weeks. Among these, 127 (43%) reported having more than one sexual partner and 135 (42%) having engaged in transactional sex. Only 16% (n=59) of confirmed cases reported contact with wild animals through hunting, handling, or consumption.

Comparing people with suspected mpox who tested positive with those who tested negative, we observed an older age distribution (median age 22 *vs* 17, p=0·034), and a higher proportion of sex workers (although statistically not significant, 13% *vs* 5%, p=0·117) and mine workers (24% *vs* 8%, p=0·005) among the confirmed cases. Compared with non-infected individuals, confirmed cases more often reported having sexual intercourse (84% *vs* 57%, p=0·001), including transactional sex (42% *vs* 16%, p=0·003), and more often reported contact with another mpox case in the three weeks preceding their diagnosis. (59% *vs* 41%, p=0·015).

### Medical History and Clinical Presentation at Admission

Only 18 (4%) mpox patients reported comorbidities, including malnutrition (n=10), HIV (n=6), tuberculosis (n=1) and chronic liver disease (n=1). (Table 2) Seven (2%) MPXV-infected participants reported smallpox vaccination during childhood. Twenty-one infected women (10%) were pregnant at the time of inclusion.

**Table 2:** Medical history and clinical presentation of confirmed mpox cases in Kamituga, DRC (2024) in relation to age categories

|  | All confirmed cases<br>(n = 431) | Per age category (years) |  |  |
| --- | --- | --- | --- | --- |
|  |  | 0-4<br>(n = 63) | 5-14<br>(n = 32) | ≥15<br>(n = 336) |
| **Medical history** | .. | .. | .. | .. |
| Childhood smallpox vaccination – n (%) | 7/308 (2%) | .. | .. | 7/308 (2%) |
| Previous mpox* – n (%) | 2/420 (1%) | 0/62 (0%) | 0/32 (0%) | 2/326 (1%) |
| Comorbidities – n (%) | 18/422 (4%) | 0/61 (0%) | 2/32 (6%) | 16/329 (5%) |
| HIV | 6/422 (1%) | 0/61 (0%) | 1/32 (3%) | 5/329 (2%) |
| Under ART | 3/6 (60%) | .. | 1/1 (100%) | 2/4 (50%) |
| Tuberculosis | 5/431 (1%) | 0/63 (0%) | 1/32 (3%) | 4/336 (1%) |
| Diabetes | 1/431 (0%) | 0/63 (0%) | 0/32 (0%) | 1/336 (0%) |
| Chronic liver disease | 1/431 (0%) | 0/63 (0%) | 1/32 (3.1) | 0/336 (0%) |
| Chronic pulmonary disease | 3/431 (1%) | 0/63 (0%) | 0/32 (0%) | 3/336 (1%) |
| Rheumatic disease | 1/431 (0%) | 0/63 (0%) | 0/32 (0%) | 1/336 (0%) |
| Arterial hypertension | 2/431 (1%) | 0/63 (0%) | 0/32 (0%) | 2/336 (1%) |
| Malnutrition# – n (%) | 10/375 (3%) | 5/56 (9%) | 3/28 (11%) | 2/291 (1%) |
| Moderate malnutrition | 9/375 (2%) | 4/56 (7%) | 3/28 (11%) | 2/291 (1%) |
| Severe malnutrition | 1/375 (0%) | 1/56 (2%) | 0/28 (0%) | 0/291 (0%) |
| Pregnancy – n (%) | 21/205 (10%) | 0 (0%) | 0 (0%) | 21/161 (13%) |
| 1st trimester | 5/20 (25%) | .. | .. | 5/20 (25%) |
| 2nd trimester | 10/20 (50%) | .. | .. | 10/20 (50%) |
| 3rd trimester | 5/20 (25%) | .. | .. | 5/20 (25%) |
| **Clinical presentation** | .. | .. | .. | .. |
| Symptoms onset, days – median (IQR) | .. | .. | .. | .. |
| Any symptom | 7 (5-10) | 6 (3-8) | 5 (4-7) | 8 (6-10) |
| Lesions | 5 (4-7) | 4 (3-6) | 3 (3-4) | 5 (4-7) |
| Prodromal phase<sup>‡</sup> – n (%) | 346/395 (88%) | 40/52 (77%) | 23/29 (79%) | 283/314 (90%) |
| Symptoms – n (%) | .. | .. | .. | .. |
| Fatigue | 384/429 (90%) | 53/62 (86%) | 28/32 (88%) | 303/335 (90%) |
| Malaise | 320/428 (75%) | 34/61 (56%) | 26/32 (81%) | 260/335 (78%) |
| Myalgia | 301/423 (71%) | 31/57 (54%) | 21/31 (68%) | 249/335 (74%) |
| Pain associated with lesions | 333/426 (78%) | 40/59 (68%) | 26/32 (81%) | 267/335 (80%) |
| Itching | 390/426 (92%) | 52/59 (88%) | 31/32 (97%) | 307/335 (92%) |
| Cough | 150/427 (35%) | 30/61 (49%) | 9/32 (28%) | 111/334 (33%) |
| Shortness of breath | 59/428 (14%) | 12/61 (20%) | 0/32 (0%) | 47/335 (14%) |
| Anorexia | 244/429 (57%) | 22/62 (36%) | 22/32 (69%) | 200/335 (60%) |
| Throat ache / dysphagia | 233/426 (55%) | 11/59 (19%) | 23/32 (72%) | 199/335 (59%) |
| Abdominal pain | 128/424 (30%) | 7/58 (12%) | 10/32 (31%) | 111/334 (33%) |
| Nausea / vomiting | 97/428 (23%) | 7/61 (12%) | 9/32 (28%) | 81/335 (24%) |
| Diarrhea | 54/428 (13%) | 7/61 (12%) | 3/32 (9%) | 44/335 (13%) |
| Headache | 249/422 (59%) | 2/56 (4%) | 18/32 (56%) | 229/334 (69%) |
| Confusion | 12/428 (3%) | 1/61 (2%) | 2/32 (6%) | 9/335 (3%) |
| Convulsions | 1/429 (0%) | 1/62 (2%) | 0/32 (0%) | 0/335 (0%) |
| Eye pain | 85/426 (20%) | 0/60 (0%) | 4/32 (13%) | 81/334 (24%) |
| Visual problems | 42/426 (10%) | 0/60 (0%) | 0/31 (0%) | 42/335 (13%) |
| Rectal pain | 46/424 (11%) | 1/59 (2%) | 0/31 (0%) | 45/334 (14%) |
| Dysuria | 168/425 (40%) | 1/58 (2%) | 1/32 (3%) | 166/335 (50%) |
| Hematuria | 22/428 (5%) | 1/61 (2%) | 0/32 (0%) | 21/335 (6%) |
| Genital pain | 117/229 (51%) | 1/58 (2%) | 0/32 (0%) | 116/221 (53%) |
| Vital signs – n (%) | .. | .. | .. | .. |
| Temperature > 38 °C | 21/424 (5%) | 4/62 (7%) | 2/32 (6%) | 15/330 (5%) |
| Tachycardia<sup>¶</sup> | 59/416 (14%) | 5/56 (9%) | 13/31 (42%) | 41/329 (13%) |
| Tachypnea<sup>¶</sup> | 110/365 (30%) | 23/54 (43%) | 12/25 (48%) | 75/286 (26%) |
| MAP < 60 mmHg | 5/397 (1%) | 2/35 (6%) | 2/31 (7%) | 1/331 (0%) |
| Oxygen saturation < 94% | 9/417 (2%) | 2/61 (3%) | 1/32 (3%) | 6/324 (2%) |
| Capillary refill > 2 sec | 18/400 (5%) | 3/59 (5%) | 3/28 (11%) | 12/313 (4%) |
| Skin lesions – n (%) | 417/428 (97%) | 59/62 (95%) | 32/32 (100%) | 326/334 (98%) |
| Active lesions<sup>£</sup> | 414/431 (96%) | 58/63 (92%) | 32/32 (100%) | 324/336 (96%) |
| Rash pattern | .. | .. | .. | .. |
| Generalized<sup>£</sup> | 141/414 (34%) | 14/58 (24%) | 11/32 (34%) | 116/324 (36%) |
| Predominantly genital<sup>§</sup> | 89/413 (22%) | 1/58 (2%) | 1/32 (3%) | 87/323 (27%) |
| Only genital | 37/414 (9%) | 1/58 (2%) | 0/32 (0%) | 36/324 (11%) |
| Lesion distribution | .. | .. | .. | .. |
| Head and neck | 298/431 (78%) | 48/63 (83%) | 26/32 (81%) | 224/336 (69%) |
| Chest | 228/431 (61%) | 31/63 (53%) | 24/32 (75%) | 173/336 (53%) |
| Abdomen | 241/431 (62%) | 36/63 (62%) | 22/32 (69%) | 183/336 (57%) |
| Back | 270/431 (71%) | 46/63 (79%) | 23/32 (72%) | 201/336 (62%) |
| Arms | 329/431 (83%) | 48/63 (83%) | 28/32 (88%) | 253/336 (78%) |
| Palms of the hands | 167/431 (42%) | 19/63 (33%) | 17/32 (53%) | 131/336 (40%) |
| Legs | 288/431 (74%) | 43/63 (74%) | 26/32 (81%) | 219/336 (68%) |
| Soles of the feet | 100/431 (20%) | 11/63 (19%) | 5/32 (16%) | 84/336 (26%) |
| Genital | 328/431 (61%) | 17/63 (29%) | 20/32 (63%) | 291/336 (90%) |
| Anal | 136/431 (29%) | 14/63 (25%) | 9/32 (28%) | 113/336 (35%) |
| Type | .. | .. | .. | .. |
| Macule | 62/414 (15%) | 15/58 (26%) | 4/32 (13%) | 43/324 (13%) |
| Papule | 222/414 (54%) | 42/58 (72%) | 24/32 (75%) | 156/324 (48%) |
| Vesicle | 291/414 (70%) | 27/58 (47%) | 15/32 (47%) | 249/324 (77%) |
| Small pustule | 178/414 (43%) | 23/58 (40%) | 11/32 (34%) | 144/324 (44%) |
| Umbilicated pustule | 156/414 (38%) | 6/58 (10%) | 8/32 (25%) | 142/324 (44%) |
| Ulcer | 94/414 (23%) | 2/58 (3%) | 3/32 (9%) | 89/324 (28%) |
| Crust | 162/414 (39%) | 15/58 (26%) | 5/32 (16%) | 142/324 (44%) |
| Scar | 51/414 (12%) | 5/58 (9%) | 2/32 (6%) | 44/324 (14%) |
| Monomorphic<sup>§</sup> | 88/414 (21%) | 22/58 (38%) | 14/32 (44%) | 52/324 (16%) |
| Complex lesions<sup>¶</sup> | 98/411 (24%) | 1/58 (2%) | 3/32 (9%) | 94/321 (29%) |
| Total lesion count – median (IQR) | 42 (17-94) | 50 (30-108) | 103 (27-186) | 38 (16-83) |
| Mucosal lesions – n (%) | 338/414 (82%) | 25/58 (43%) | 26/32 (81%) | 287/324 (89%) |
| Oral | 102/412 (25%) | 11/58 (19%) | 12/32 (38%) | 79/322 (25%) |
| Tonsillar | 187/410 (46%) | 13/57 (23%) | 19/32 (59%) | 155/321 (48%) |
| Penile | 149/215 (69%) | 9/31 (29%) | 6/16 (38%) | 134/168 (80%) |
| Vaginal | 144/195 (74%) | 6/26 (23%) | 8/15 (53%) | 130/154 (84%) |
| Lymphadenopathy – n (%) | 295/414 (71%) | 9/58 (16%) | 22/32 (69%) | 264/324 (82%) |
| Retro-auricular | 48/414 (12%) | 5/58 (9%) | 8/32 (25%) | 35/324 (11%) |
| Submandibular | 90/414 (22%) | 8/58 (14%) | 15/32 (47%) | 67/324 (21%) |
| Cervical | 27/414 (7%) | 1/58 (2%) | 2/32 (6%) | 24/324 (7%) |
| Supra clavicular | 2/414 (1%) | 0/58 (0%) | 0/32 (0%) | 2/324 (1%) |
| Axillar | 23/414 (6%) | 0/58 (0%) | 1/32 (3%) | 22/324 (7%) |
| Inguinal | 260/414 (63%) | 4/58 (7%) | 10/32 (31%) | 246/324 (76%) |

Mpox-confirmed patients presented at the study site a median of seven (IQR 5 – 10) days after the onset of symptoms and five (IQR 4-7) days after the appearance of skin lesions (Table 2). Prodromal symptoms preceded the rash in 346 (88%) of patients. Systemic symptoms such as fatigue, malaise and myalgia were highly prevalent, affecting 384 (90%), 320 (75%) and 301 (71%) of patients, respectively. The lesions commonly caused itching (92%) or pain (78%). Eye pain and visual problems were reported in 20% and 10% of cases respectively, mostly among adults. Adult mpox patients often had genital pain (n=116, 53%), dysuria (n=166, 50%), or rectal pain (n=45, 14%). Anogenital symptoms rarely occurred among children.

At inclusion, 21 (5%) of mpox cases had a temperature above 38°C (Table 2). Active skin lesions were present in 414 (96%) patients. The median total lesion count at admission was 42 (IQR 17-94, range 1-1420). Most participants (n=326, 79%) had a combination of different lesion types, including vesicles (n=291, 70%), papules (n= 222, 54%) and small pustules (n=178, 43%). Umbilicated pustules were observed in 156 participants (38%), ulcers in 94 (23%) and macules in 62 (15%). Children up to 14 years old, however, mostly presented with papules (n=66, 73%) and umbilicated pustules were rare (n=14, 16%).

One third (n=141, 34%) of patients had generalized rash, defined as lesions on at least eight out of ten body regions (Table 2). Less than half of patients had lesions on the palms of the hands (n=167, 42%) or on the soles of the feet (n=100, 20%). Adults more often had genital skin lesions than children (90% *vs* 41%, p<0·001). (Figure 1B, Table 2) The genital area made up > 50% of total lesion count in 27% of adults and was the only localization of lesions in 11% of adults (*vs* 2 and 1%, respectively, in children). Among adults, the genital area had the highest lesion density, while in children, lesions were more concentrated on the abdomen, thorax, arms, and genital area (Figure 1C).

Complex lesions, defined as large (> 2cm in diameter), confluent or necrotic lesions were present in 98 (24%) mostly adult patients (n=94), of whom 93 (95%) reported these lesions in the genital region (Table 2). Mucosal lesions (n=338, 82% for any mucosal lesion) were common across age groups, although genital mucosal lesions were more prevalent in adults compared to children up to 14 years old (82% *vs* 33%, p<0·001). Lymphadenopathy differed significantly over age groups (p<0·001). While common in adults (n=264, 82%) and children aged 5-14 years (n=22, 69%), lymphadenopathy was less frequent in children under five years old (n=9, 16%). In adults, the inguinal nodes were primarily affected, whereas in children, the submandibular nodes were more commonly involved.

### Outcome and complications during hospitalization

Of the 431 MPXV-infected participants, all but four confirmed participants (99%) accepted to be hospitalized, of which 17 discharged themselves against medical advice. Two children under five years old died during hospitalization, giving an in-hospital mortality 0·5% (95% CI [0·1%, 1·7%]).

Detailed longitudinal follow-up data were available for 315 (73%) hospitalized patients. (Table 3) Among those, the median duration of hospitalization was six (IQR 5-9) days. The total lesion count increased during hospitalization in 148 (34·3%) participants. The median maximum lesion count was 59 (IQR 23-139, range 1-1420) and occurred a median of 7 days (IQR 5-9) after the onset of lesions. Based on maximum lesion count and according to the WHO clinical severity score, 78 (25%) patients had mild disease (<25 lesions), 127 (41%) had moderate disease (25-99 lesions), 60 (19%) had severe disease (100-249 lesions) and 47 (15%) had grave disease. The median time from onset to resolution of lesions was 14 (95% CI 14-16) days, and of symptoms overall was 18 (95% CI 17-19) days. (Figure 1E and F)

**Table 3.**
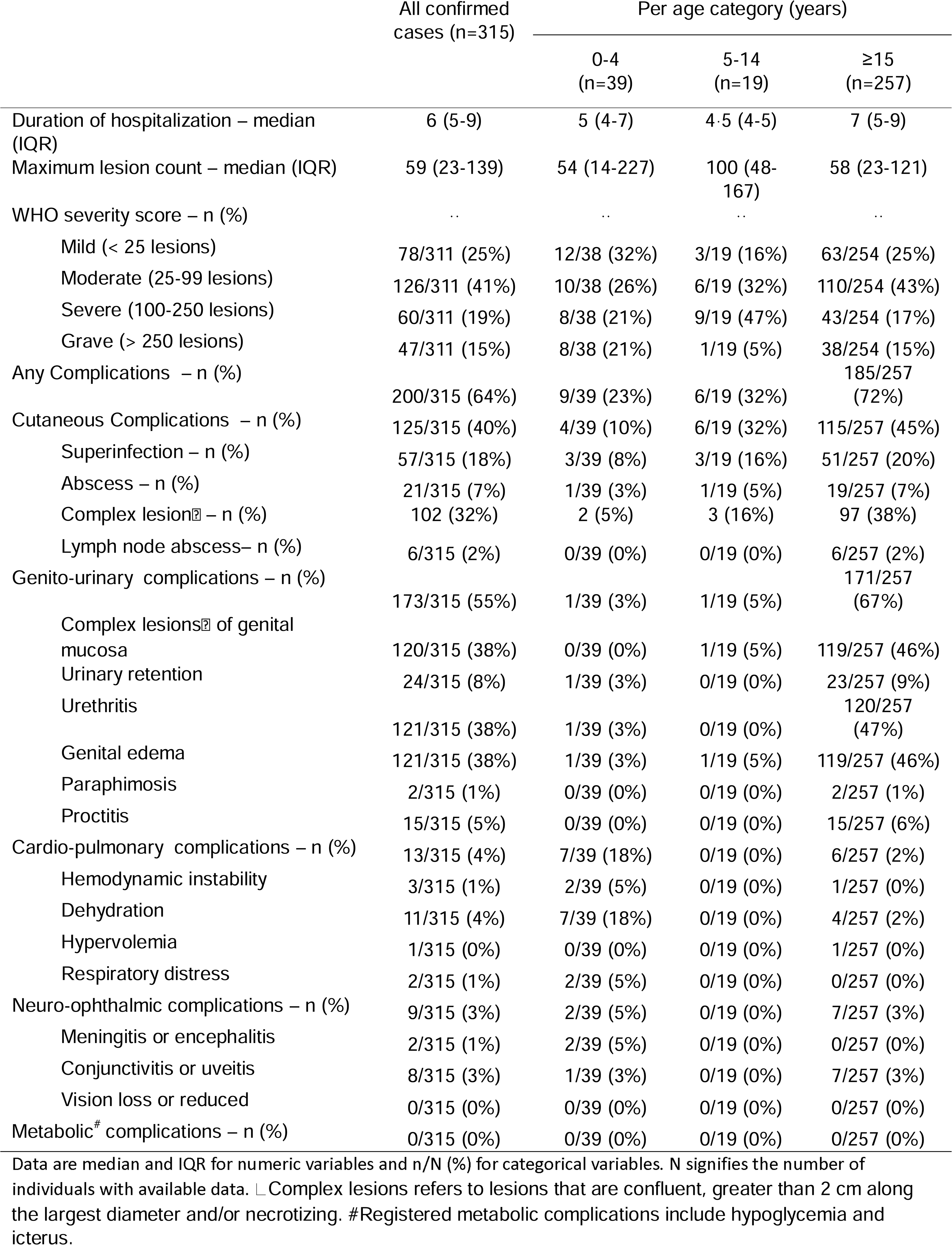
Complications at any timepoint during hospitalization for mpox treatment in Kamituga, DRC (2024)

|  | All confirmed cases (n=315) | Per age category (years) |  |  |
| --- | --- | --- | --- | --- |
|  |  | 0-4 (n=39) | 5-14 (n=19) | ≥15 (n=257) |
| Duration of hospitalization – median (IQR) | 6 (5-9) | 5 (4-7) | 4.5 (4-5) | 7 (5-9) |
| Maximum lesion count – median (IQR) | 59 (23-139) | 54 (14-227) | 100 (48-167) | 58 (23-121) |
| WHO severity score – n (%) | .. | .. | .. | .. |
| Mild (< 25 lesions) | 78/311 (25%) | 12/38 (32%) | 3/19 (16%) | 63/254 (25%) |
| Moderate (25-99 lesions) | 126/311 (41%) | 10/38 (26%) | 6/19 (32%) | 110/254 (43%) |
| Severe (100-250 lesions) | 60/311 (19%) | 8/38 (21%) | 9/19 (47%) | 43/254 (17%) |
| Grave (> 250 lesions) | 47/311 (15%) | 8/38 (21%) | 1/19 (5%) | 38/254 (15%) |
| Any Complications – n (%) | 200/315 (64%) | 9/39 (23%) | 6/19 (32%) | 185/257 (72%) |
| Cutaneous Complications – n (%) | 125/315 (40%) | 4/39 (10%) | 6/19 (32%) | 115/257 (45%) |
| Superinfection – n (%) | 57/315 (18%) | 3/39 (8%) | 3/19 (16%) | 51/257 (20%) |
| Abscess – n (%) | 21/315 (7%) | 1/39 (3%) | 1/19 (5%) | 19/257 (7%) |
| Complex lesion<sup>‡</sup> – n (%) | 102 (32%) | 2 (5%) | 3 (16%) | 97 (38%) |
| Lymph node abscess– n (%) | 6/315 (2%) | 0/39 (0%) | 0/19 (0%) | 6/257 (2%) |
| Genito-urinary complications – n (%) | 173/315 (55%) | 1/39 (3%) | 1/19 (5%) | 171/257 (67%) |
| Complex lesions<sup>‡</sup> of genital mucosa | 120/315 (38%) | 0/39 (0%) | 1/19 (5%) | 119/257 (46%) |
| Urinary retention | 24/315 (8%) | 1/39 (3%) | 0/19 (0%) | 23/257 (9%) |
| Urethritis | 121/315 (38%) | 1/39 (3%) | 0/19 (0%) | 120/257 (47%) |
| Genital edema | 121/315 (38%) | 1/39 (3%) | 1/19 (5%) | 119/257 (46%) |
| Paraphimosis | 2/315 (1%) | 0/39 (0%) | 0/19 (0%) | 2/257 (1%) |
| Proctitis | 15/315 (5%) | 0/39 (0%) | 0/19 (0%) | 15/257 (6%) |
| Cardio-pulmonary complications – n (%) | 13/315 (4%) | 7/39 (18%) | 0/19 (0%) | 6/257 (2%) |
| Hemodynamic instability | 3/315 (1%) | 2/39 (5%) | 0/19 (0%) | 1/257 (0%) |
| Dehydration | 11/315 (4%) | 7/39 (18%) | 0/19 (0%) | 4/257 (2%) |
| Hypervolemia | 1/315 (0%) | 0/39 (0%) | 0/19 (0%) | 1/257 (0%) |
| Respiratory distress | 2/315 (1%) | 2/39 (5%) | 0/19 (0%) | 0/257 (0%) |
| Neuro-ophthalmic complications – n (%) | 9/315 (3%) | 2/39 (5%) | 0/19 (0%) | 7/257 (3%) |
| Meningitis or encephalitis | 2/315 (1%) | 2/39 (5%) | 0/19 (0%) | 0/257 (0%) |
| Conjunctivitis or uveitis | 8/315 (3%) | 1/39 (3%) | 0/19 (0%) | 7/257 (3%) |
| Vision loss or reduced | 0/315 (0%) | 0/39 (0%) | 0/19 (0%) | 0/257 (0%) |
| Metabolic<sup>#</sup> complications – n (%) | 0/315 (0%) | 0/39 (0%) | 0/19 (0%) | 0/257 (0%) |

Data are median and IQR for numeric variables and n/N (%) for categorical variables. N signifies the number of individuals with available data. <sup>‡</sup> Complex lesions refers to lesions that are confluent, greater than 2 cm along the largest diameter and/or necrotizing. <sup>#</sup>Registered metabolic complications include hypoglycemia and icterus.

Overall, 200 (64%) of the 315 hospitalized patients experienced complications during observation, including genital oedema or urethritis (n=121, 38%, each), complex mucosal lesions (n=120, 38%), complex skin lesions (n=102, 32%), secondary bacterial infections (n=57, 18%) or abscess formation (n=21, 7%) (Table 3). Other complications were rare (< 10%).

### Persistent symptoms at follow-up

In total, 208 cases (43%) returned for assessment on day 29 and 103 cases (24%) on day 59. (Table 4) At follow-up on day 29, 2 (1%), participants reported a decline in general health compared to before the onset of mpox. No one reported a decline in general health on day 59.

**Table 4.** Clinical sequelae at day 29 and day 59 after mpox diagnosis in Kamituga, DRC (2024)

|  | Day 29 (n =208) | Day 59 (n = 102) |
| --- | --- | --- |
| Deterioration of general health since mpox – n (%) | 2/206 (1%) | 0/102 (0%) |
| Medical care for ongoing health issues – n (%) | 11/81 (14%) | 3/38 (8%) |
| **Persistent symptoms** | .. | .. |
| Any – n (%) | 78/208 (38%) | 38/103 (37%) |
| Constitutional – n (%) | 20/208 (10%) | 9/103 (9%) |
| Fatigue | 9/207 (4%) | 4/103 (4%) |
| Malaise | 5/207 (2%) | 0/103 (0%) |
| Myalgia | 6/207 (3%) | 0/103 (0%) |
| Headache | 11/206 (5%) | 5/100 (5%) |
| Anorexia | 2/208 (1%) | 2/102 (2%) |
| Skin – n (%) | 40/208 (19%) | 15/102 (15%) |
| Pain associated with lesions | 5/208 (2%) | 0/102 (0%) |
| Itching | 36/207 (17%) | 15/101 (15%) |
| Psychiatric – n (%) | 13/208 (6%) | 6/103 (6%) |
| Depression | 1/208 (1%) | 0/103 (0%) |
| Anxiety | 13/208 (6%) | 6/102 (6%) |
| Respiratory – n (%) | 9/208 (4%) | 6/103 (6%) |
| Cough | 7/208 (3%) | 6/103 (6%) |
| Shortness of breath | 2/208 (1%) | 0/103 (0%) |
| Gastro-intestinal – n (%) | 11/208 (5%) | 2/103 (2%) |
| Throat ache / dysphagia | 5/208 (2%) | 0/102 (0%) |
| Abdominal pain | 5/206 (2%) | 1/101 (1%) |
| Nausea / vomiting | 2/208 (1%) | 1/103 (1%) |
| Diarrhea | 3/208 (1%) | 0/103 (0%) |
| Rectal pain | 1/208 (1%) | 0/103 (0%) |
| Pain on defecation | 2/207 (1%) | 0/103 (0%) |
| Genito-urinary – n (%) | 15/208 (7%) | 4/103 (4%) |
| Dysuria | 5/208 (2%) | 0/103 (0%) |
| Genital pain | 3/172 (2%) | 1/87 (1%) |
| Dyspareunia | 4/159 (3%) | 2/86 (2%) |
| Vaginal discharge | 4/84 (5%) | 2/41 (5%) |
| Vaginal dryness | 6/82 (7%) | 4/41 (10%) |
| Adhesions of the foreskin | 1/107 (1%) | 0/52 (0%) |
| Painful erection | 2/87 (2%) | 0/45 (0%) |
| Eye – n (%) | 22/207 (11%) | 18/102 (18%) |
| Eye pain | 9/204 (4%) | 4/101 (4%) |
| Photophobia | 14/203 (7%) | 11/100 (11%) |
| Itching | 15/206 (7%) | 12/102 (12%) |
| Foreign body sensation | 8/203 (4%) | 9/99 (9%) |
| Reduced visual acuity | 5/206 (2%) | 5/101 (5%) |
| Blindness | 0/207 (0%) | 1/101 (1%) |
| **Persistent lesions** | .. | .. |
| Active skin lesions – n (%) | 5/206 (2%) | 1/103 (1%) |
| Scars – n (%) | 153/208 (74%) | 51/103 (50%) |
| Eye – n (%) | 9/208 (4%) | 2/103 (2%) |
| Eye lid edema | 1/207 (1%) | 1/102 (1%) |
| Palpebral lesion | 1/207 (1%) | 1/102 (1%) |
| Conjunctival lesion | 1/207 (1%) | 2/102 (2%) |
| Conjunctivitis / uveitis | 8/207 (4%) | 2/102 (2%) |

Data are n/N (%), with N representing the number of individuals with available data.

A total of 78 (38%) participants experienced residual symptoms on day 29, and 38 (37%) on day 59. Skin-related symptoms were the most common, affecting 40 (19%) participants on day 29 and 15 (15%) on day 59. Eye symptoms were reported by 22 (11%) participants on day 29 and 18 (18%) on day 59. Five participants described reduced visual acuity on day 29 (2%) and day 59 (5%), and one person reported blindness on day 59 (1%). Other symptoms occurred in < 10% of participants on days 29 and 59. (Table 4)

On day 29, 153 (74%) participants had scars and five (2%) had active skin lesions in the form of ulcers. However, upon follow-up at day 59, only 51 (50%) still had discernible scars (Figure 1D), and one (1%) participant still had an unresolved genital ulcer. Residual mucosal lesions were noted in six (3%) participants on day 29 and one participant (1%) on day 59. (Table 4)

### Pregnancy outcomes

Among the 21 pregnant women in the cohort, no adverse pregnancy outcomes occurred during hospitalization. Following the initial admission, the final pregnancy outcomes were recorded for six participants, where 4/6 (67%) experienced an adverse pregnancy outcome. Two women suffered spontaneous abortions, during first trimester and second trimester, respectively. Additionally, two women showed evidence of vertical transmission of MPXV to their neonates. One fetus died in utero near the end of the second trimester, requiring induction of labor. Mpox skin lesions were discernible on the head upon delivery. The other child was born full term via Caesarean section due to prolonged labor complicated by uterine rupture. This infant also presented with characteristic mpox skin lesions, suffered from asphyxia, and died shortly after birth. The remaining two women delivered healthy full-term babies.

## Discussion

This study provides a prospective, detailed characterization of patient profiles in the emerging Clade Ib outbreak. Reflecting the evolving transmission pattern of Clade Ib, mpox predominantly affected adolescents and young adults, though not exclusively. Exposure data indicate that sexual contact is a significant driver of transmission within this group, as corroborated by the prevalent genital presentations among adults. Most patients experience mild to moderate disease, with complications that are primarily genito-urinary in nature, and clinically significant sequelae other than scars are rare. Approximately one-fifth of cases, however, were children, who typically present with more extragenital disease.

This study was conducted in a remote area of Eastern DRC, where access to high-quality healthcare is limited. However, through collaboration with the health authorities, and humanitarian organizations, we enhanced the treatment facilities and level of care provided to mpox patients. This enabled the study to be conducted within an ethically sound framework and created a suitable environment for high-quality data collection.

Nevertheless, working in resource-limited settings comes with inherent limitations. Firstly, while the on-site diagnostic platform allowed for the detection of OPXV DNA, point-of-care Clade-specific confirmation is currently unavailable. Given that Kamituga is situated at the original epicenter of the Clade Ib outbreak and has no history of Clade Ia outbreaks, it is highly probable that all study participants are infected with Clade Ib. At the time of this preprint’s publication, Clade confirmation is underway for a subset of participants. Secondly, despite implementing a broad testing strategy, recruitment relied on passive case detection, and the PCR positivity rate remained consistently high throughout the study (around 80%). This suggests that a significant proportion of cases in the community may have gone undetected. If these undetected cases involve individuals with atypical presentations, very mild symptoms, or severe illness (including community deaths), their absence from the study could introduce bias into the results. Additionally, the study had a large sample size, but there was substantial loss to follow-up in the consecutive visits, which could impact the representativeness of the data for the long-term outcomes. Finally, resource limitations restricted our ability to perform additional diagnostic testing, including for HIV, other sexually transmitted infections, Varicella, hematological and biochemical analyses, as well as assessments for complications like bacterial superinfections and sepsis.

Despite these limitations, our findings allow for a comprehensive comparison between this Clade Ib outbreak and other mpox outbreaks. While Clade Ia outbreaks in endemic regions in the DRC are typically driven by zoonotic spillover,^6^ exposure to wild animals - and thus a potential animal reservoir - was rare in our cohort. In contrast, 59% of patients recalled contact with an index case, of which 42% included sexual contact. Additionally, the overrepresentation of occupations such as sex workers and mine workers (who make up most of their clientele) and the higher levels of sexual activity and more frequent transactional sex among PCR-positive mpox cases compared to PCR-negative suspects, point toward sexual contact as one of the drivers of transmission, similar to patterns observed in Clade IIb.^4^ However, in contrast to the global Clade IIb outbreak,^4^ the equal gender distribution among mpox cases in our study suggests that transmission predominantly occurred through heterosexual contact rather than male-to-male sexual contact. However, because of cultural sensitivities, we did not collect information on the gender of sexual partners.

Similar to Clade IIb infections, distinct anogenital manifestations were observed in adults with Clade Ib mpox.^1,4,17^ Up to one-third of adults presented with predominantly genital lesions, and over three-quarters exhibited involvement of the genital mucosa and/or inguinal lymphadenopathy. In contrast, most previous Clade Ia studies reported genital lesions in less than 30% of cases, typically showing a more generalized lesion distribution.^8,18–20^ These findings suggest that the mode of MPXV transmission might determine disease presentation. Interestingly, despite similarities in disease presentation between Clades Ib and IIb, Clade Ib infections in adults appear to be somewhat more severe. While only 12% of Clade IIb patients had over 20 lesions and 0–4% had more than 100, in our cohort, 66% had more than 25 lesions, and 24% exceeded 100 lesions.^1^ This could be caused by differences in the affected populations or in virulence between clades.

Conversely, children under 15 years exhibited a more typical clinical presentation featuring widespread lesions in a centrifugal distribution, though with a low frequency of lesions on the palms and soles.^18–22^ In contrast to Clade Ia outbreaks, where they typically make up three-quarters of cases,^5,8,20^ children represented only a minority of cases in our study. Transmission dynamics in children remain unclear; however, our data suggest that they were primarily infected within the household. Household transmission was rarely reported during the global Clade IIb outbreak, although in a Nigerian cohort up to 16% were children under 18 years old.^23^ Moreover, in other health zones in South Kivu affected by the Clade Ib outbreak and parts of neighboring Burundi, children constitute the majority of confirmed mpox cases.^2,24^ Since our study was limited to a single health zone, caution is advised in generalizing these findings. Further studies on transmission dynamics are essential.

A final important point of comparison with previously documented outbreaks are the outcomes and complications of mpox. Long-term follow-up revealed few severe sequelae other than scars among Clade Ib infected individuals, although one patient suffered from blindness.^18,19^ Of particular concern though, are adverse pregnancy outcomes; in our study, four out of six women with recorded pregnancy outcomes lost their child, consistent with findings from Clade Ia.^7,25^ In Clade IIb, no cases of presumed or confirmed vertical transmission have been reported.^26^ To better understand the impact of MPXV infection on fetal development and perinatal morbidity and mortality, larger prospective cohorts of pregnant women are essential.

In-hospital mortality in our cohort was low at 0·5%, with all deaths recorded in children under five. This mortality rate is higher than the 0·2% observed during the global Clade IIb outbreak in 2022—though rates as high as 6% have been reported in Nigeria Click or tap here to enter text.^23^ — but lower than the 1·4% to 17% reported for Clade Ia.^7,10,18,19,27,28^ Factors such as viral virulence, transmission mode, reporting bias, population age structure, underlying comorbidities, and healthcare access may all contribute to these observed differences in mortality.

This study provides a comprehensive overview of the Clade Ib outbreak in South Kivu, highlighting shifts in transmission dynamics, clinical presentation, and patient outcomes that differ from previous Clade I and II outbreaks. Future public health interventions – such as enhanced surveillance, integrated patient management, infection control policies, contact tracing and other targeted active case detection strategies – should reflect these evolving patterns of mpox presentation. Our findings will also guide and help prioritize further research initiatives.

## Data Availability

All data produced in the present study are available upon reasonable request to the authors

## Acknowledgments

We thank all patients who agreed to be included in this cohort and particularly those who provided additional consent for publication of their deidentified images. We acknowledge the dedicated staff of KGH and ALIMA for their continuing expert patient care. We also acknowledge the support of the Health Zone and the Provincial Health Department to facilitate setting up of the study and the KGH for generously disposing the study team of necessary infrastructure to pursue study activities. Finally, we thank our colleagues Inge Van Cauwenberg, Marie Stéphanie Smet, Lydia Lunda, Alliance Mbandu, and Plamedi Fatouma from ITM Kinshasa office administrative and logistical support.

## Author contributions

Conceptualized the study and wrote the protocol: IB, EHV, EDV, EB, YM, SSN, LL, PMK. Participant inclusion, investigation and data collection: GM, PM, JCT, LMM, FMK. Conducted data analysis: IB, EHV, EDV, AT, CVD, SSN, LL. Data management: SH, YM. Secured funding: PKM, LL, JJMT, DM, EBM, PK, NL, AR, JA. Supervision study proceedings: LMM, FMK, CK, DM, SA, JJMT, SSn, LL, PMK. Supervised laboratory procedures: JCT, EKL, JCMC, TW, KV, DM, SA. Writing of the initial manuscript: IB, EHV, NL, SSN, LL, PMK. Reviewed the manuscript: IB, EHV, GM, PM, JCT, EDV, EB, YM, AT, CVD, EA, LMM, FMK, SH, NH, EKL, JCMC, AR, JK, NL, PK, EBM, JA, OT, RK, KV, TW, CK, DM, SA, JJMT, SSN, LL, PMK. Had full access to the data: IB, EHV, AT, SSN, LL, PMK. All authors contributed to review and approved the final version of the manuscript.

## Data sharing

De-identified participant data collected for the study will be made available from the corresponding author upon reasonable request (i.e., when ethically viable without violating the protection of participants or other valid ethical, privacy, or security concerns).

## References

1. Mitjà O, Ogoina D, Titanji BK, Galvan C, Muyembe JJ, Marks M, et al. Monkeypox. The Lancet. 2023 Jan 7;401(10370):60–74.

2. 2022-24 Mpox (Monkeypox) Outbreak: Global Trends [Internet]. [cited 2024 Sep 24]. Available from: https://worldhealthorg.shinyapps.io/mpx_global/#43_Case_profile_(overall)

3. Yinka-Ogunleye A, Aruna O, Dalhat M, Ogoina D, McCollum A, Disu Y, et al. Outbreak of human monkeypox in Nigeria in 2017–18: a clinical and epidemiological report. Lancet Infect Dis. 2019 Aug 1;19(8):872.

4. Thornhill JP, Barkati S, Walmsley S, Rockstroh J, Antinori A, Harrison LB, et al. Monkeypox Virus Infection in Humans across 16 Countries - April-June 2022. N Engl J Med. 2022 Aug 25;387(8):679–91.

5. Bangwen E, Diavita R, De Vos E, Hasivirwe Vakaniaki E, Nundu SS, Mutombo A, et al. Mpox in the Democratic Republic of Congo: Analysis of National Epidemiological and Laboratory Surveillance Data, 2010 - 2023. 2024 [cited 2024 Oct 23]; Available from: https://papers.ssrn.com/abstract=4954317

6. Kinganda-Lusamaki E, Amuri-Aziza A, Fernandez-Nuñez N, Makangara-Cigolo JC, Pratt C, Vakaniaki EH, et al. Clade I mpox virus genomic diversity in the Democratic Republic of the Congo, 2018–2024: Predominance of zoonotic transmission. Cell. 2024 Oct;0(0).

7. Pittman PR, Martin JW, Kingebeni PM, Tamfum JJM, Mwema G, Wan Q, et al. Clinical characterization and placental pathology of mpox infection in hospitalized patients in the Democratic Republic of the Congo. PLoS Negl Trop Dis. 2023 Apr 1;17(4).

8. Whitehouse ER, Bonwitt J, Hughes CM, Lushima RS, Likafi T, Nguete B, et al. Clinical and Epidemiological Findings from Enhanced Monkeypox Surveillance in Tshuapa Province, Democratic Republic of the Congo During 2011–2015. J Infect Dis. 2021 Jun 4;223(11):1870–8.

9. McCollum AM, Nakazawa Y, Ndongala GM, Pukuta E, Karhemere S, Lushima RS, et al. Human Monkeypox in the Kivus, a Conflict Region of the Democratic Republic of the Congo. Am J Trop Med Hyg. 2015 Oct 1;93(4):718–21.

10. Vakaniaki EH, Kacita C, Kinganda-Lusamaki E, O’Toole Á, Wawina-Bokalanga T, Mukadi-Bamuleka D, et al. Sustained human outbreak of a new MPXV clade I lineage in eastern Democratic Republic of the Congo. Nat Med. 2024;

11. Masirika LM, Udahemuka JC, Schuele L, Ndishimye P, Otani S, Mbiribindi JB, et al. Ongoing mpox outbreak in Kamituga, South Kivu province, associated with monkeypox virus of a novel Clade I sub-lineage, Democratic Republic of the Congo, 2024. Eurosurveillance. 2024 Mar 14;29(11).

12. Nextstrain / groups / inrb-mpox / clade-I [Internet]. [cited 2024 Nov 7]. Available from: https://nextstrain.org/groups/inrb-mpox/clade-I?c=date_submitted&f_clade_membership=clade%20Ib&f_division=Sud%20Kivu

13. Murhula Masirika L, Claude Udahemuka J, Schuele L, Nieuwenhuijse DF, Ndishimye P, Boter M, et al. Mapping and sequencing of cases from an ongoing outbreak of Clade Ib monkeypox virus in South Kivu, Eastern Democratic Republic of the Congo between September 2023 to June 2024. medRxiv. 2024 Sep 19;2024.09.18.24313835.

14. WHO Director-General declares mpox outbreak a public health emergency of international concern [Internet]. [cited 2024 Sep 24]. Available from: https://www.who.int/news/item/14-08-2024-who-director-general-declares-mpox-outbreak-a-public-health-emergency-of-international-concern

15. Ministère de la santé publique hygiène et prévention P du SKDP de la santé. Pyramide sanitaire des zones de sante 2024. 2024.

16. Damhorst GL, McLendon K, Morales E, Solis ZM, Fitts E, Bowers HB, et al. Performance of the Xpert^TM^ Mpox PCR assay with oropharyngeal, anorectal, and cutaneous lesion swab specimens. J Clin Virol. 2024 Apr 1;171.

17. Català A, Clavo-Escribano P, Riera-Monroig J, Martín-Ezquerra G, Fernandez-Gonzalez P, Revelles-Peñas L, et al. Monkeypox outbreak in Spain: clinical and epidemiological findings in a prospective cross-sectional study of 185 cases. British Journal of Dermatology. 2022 Nov 1;187(5):765–72.

18. Ježek Z, Szczeniowski M, Paluku KM, Mutombo M. Human Monkeypox: Clinical Features of 282 Patients. J Infect Dis. 1987 Aug 1;156(2):293–8.

19. Jezek Z, Grab B, Szczeniowski M, Paluku KM, Mutombo M. Clinico-epidemiological features of monkeypox patients with an animal or human source of infection. Bull World Health Organ. 1988;66(4):459.

20. Osadebe L, Hughes CM, Shongo Lushima R, Kabamba J, Nguete B, Malekani J, et al. Enhancing case definitions for surveillance of human monkeypox in the Democratic Republic of Congo. PLoS Negl Trop Dis. 2017 Sep 11;11(9).

21. Hoff NA, Morier DS, Kisalu NK, Johnston SC, Doshi RH, Hensley LE, et al. Varicella Coinfection in Patients with Active Monkeypox in the Democratic Republic of the Congo. Ecohealth. 2017 Sep 1;14(3):564–74.

22. Hughes CM, Liu L, Davidson WB, Radford KW, Wilkins K, Monroe B, et al. A tale of two viruses: Coinfections of monkeypox and varicella zoster virus in the democratic republic of congo. American Journal of Tropical Medicine and Hygiene. 2021 Feb 1;104(2):604–11.

23. Ogoina D, Dalhat MM, Denue BA, Okowa M, Chika-Igwenyi NM, Yusuff HA, et al. Clinical characteristics and predictors of human mpox outcome during the 2022 outbreak in Nigeria: a cohort study. Lancet Infect Dis. 2023 Dec 1;23(12):1418–28.

24. Nizigiyimana A, Ndikumwenayo F, Houben S, Manirakiza M, van Lettow M, Liesenborghs L, et al. Epidemiological analysis of confirmed mpox cases, Burundi, 3 July to 9 September 2024. Euro Surveill. 2024 Oct 17;29(42).

25. Mbala PK, Huggins JW, Riu-Rovira T, Ahuka SM, Mulembakani P, Rimoin AW, et al. Maternal and Fetal Outcomes among Pregnant Women with Human Monkeypox Infection in the Democratic Republic of Congo. Journal of Infectious Diseases. 2017 Nov 1;216(7):824–8.

26. Sanchez Clemente N, Coles C, Paixao ES, Brickley EB, Whittaker E, Alfven T, et al. Paediatric, maternal, and congenital mpox: a systematic review and meta-analysis. Lancet Glob Health. 2024 Apr 1;12(4):e572–88.

27. Breman JG, Kalisa-Ruti, Steniowski M V., Zanotto E, Gromyko AI, Arita I. Human monkeypox, 1970-79. Bull World Health Organ. 1980;58(2):165.

28. Hutin YJF, Williams RJ, Malfait P, Pebody R, Loparev VN, Ropp SL, et al. Outbreak of Human Monkeypox, Democratic Republic of Congo, 1996 to 1997. Emerg Infect Dis. 2001 Jun;7(3):434–8.

