## Supplementary figures and images for "Epidemiological and Clinical Features of Mpox during the Clade Ib Outbreak in South-Kivu, Democratic Republic of the Congo: a Prospective Cohort Study"

### Supplemental Figure 1

**Supplementary figure 1. Flowchart participants**

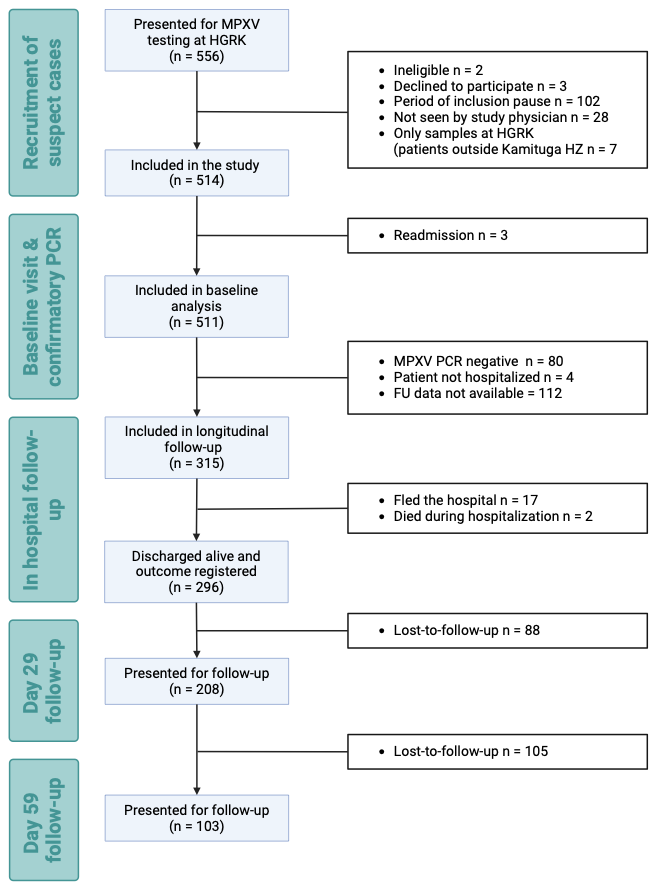
